# COPD in the time of COVID-19: An analysis of acute exacerbations and reported behavioural changes in patients with COPD

**DOI:** 10.1101/2020.09.18.20197202

**Authors:** Hamish McAuley, Kate Hadley, Omer Elneima, Christopher E Brightling, Rachael A Evans, Michael C Steiner, Neil J Greening

## Abstract

**Introduction:** The impact of the SARS-CoV-2 pandemic, and lockdown measures, on acute exacerbations of COPD (AECOPD) is unknown. We aimed to evaluate the change in AECOPD treatment frequency during the first six weeks of lockdown in the UK compared with 2019 and assess changes in self-reported behaviour and well-being.

**Methods:** In this observational study patients with established COPD were recruited. Exacerbation frequency was measured in the first six weeks of COVID lockdown and compared with the same period in 2019 using electronic health records. A telephone survey was used to assess changes in anxiety, inhaler adherence, physical activity, shopping and visitor behaviour during the pre-lockdown and lockdown periods compared to normal.

**Results:** 160 participants were recruited (mean [SD] age 67.3 [8.1] years, 88 [55%] male, FEV1 34.3 [13] % predicted) and 140 [88%] reported at least one AECOPD in the previous year. Significantly more community treated exacerbations were observed in 2020 compared with 2019 (126 vs 99, p=0.026). The increase was as a result of multiple courses of treatment, with a similar proportion of patients receiving at least one course (34.4% vs 33.8%).

During “lockdown” participants reported significantly increased anxiety, adherence to their preventative inhalers, and good adherence to shielding advice (all p<0.001). A significant reduction in self-reported physical activity and visitors was reported (both p<0.001).

**Discussion:** Treatment for AECOPD events increased during the first six weeks of the SARS-CoV-2 pandemic in the UK compared to 2019. This was associated with increased symptoms of anxiety and significant behavioural change.

**What is the key question?:** How has the COVID-19 pandemic lockdown affected exacerbation patterns and behaviour in patients with COPD?

**What is the bottom line?:** A 38% increase in the number of community treated exacerbations was seen in 2020 compared with 2019. Increased symptoms of anxiety were seen in patients, alongside increased inhaler adherence and reduced physical activity.

**Why read on?:** The impact of the COVID-19 pandemic has led to significant changes in treatment for exacerbations of COPD in the community, as well as increased anxiety amongst patients.

## INTRODUCTION

Acute Exacerbations of Chronic Obstructive Pulmonary Disease (AECOPD) are a frequent problem for people with COPD, adversely affecting morbidity and mortality and are an important cause of unscheduled healthcare contacts including admission to hospital^1^. The Global Initiative for COPD (GOLD) report grades the severity of these events according to treatment requirement, defining moderate events as those needing community provision of oral antibiotics and corticosteroids and severe events as those requiring hospitalisation^2^.

Healthcare provision for people with COPD has been impacted by the Severe Acute Respiratory Syndrome Coronavirus 2 (SARS-CoV-2) pandemic through the requirement for more distant/remote contact with the healthcare team to reduce the risk of virus transmission^3^. Additionally, because of the appreciation of a greater risk of morbidity from SARS-CoV-2 infection^4^ more stringent social isolation has been recommended for people with COPD during the period of societal lockdown that has been implemented in most countries affected by the pandemic. In the UK this has been termed “shielding” and includes advice against leaving home for any reason other than for essential work or shopping with very limited exceptions.

During this time healthcare professionals, providing care for people with COPD have reported lower than expected presentation rates for AECOPD in both community and acute hospital settings^5 6^. However, it is unclear whether this is due to a genuine reduction in AECOPD rates (potentially due to lower respiratory viral transmission^7^ and/or atmospheric pollution) or due to higher thresholds for patient reporting to healthcare services because of fearfulness about contracting SARS-CoV-2 in healthcare environments, particular hospitals. In addition there is limited understanding of the impact of enhanced shielding on the psychological wellbeing and physical activity in people with pre-existing respiratory disease such as COPD^8 9^. Having a chronic condition such as COPD does not appear to increase the likelihood of SARS-CoV-2 infection^10^, it does convey increased risk of hospitalisation and death^4^.

Firstly, in this observational study we recorded the change in moderate and severe AECOPD treatment frequency (assessed objectively through prescription records or requirement for hospitalisation) during the first six weeks of societal lockdown in the UK compared with the equivalent period 12 months previously. Secondly, we assessed self-reported behaviour change during the pre-lockdown and lockdown period by telephone interview in order to explore potential reasons for any observed changes in AECOPD treatment frequency

## METHODS

### Study Design

We compared rates of treatment for exacerbations of COPD managed in the community and hospital setting between the first six-week period of the SARS-CoV-2 “lockdown” in England (15^th^ March 2020 to 30^th^ April 2020) with the same six-week period the previous year (15^th^ March 2019 to 30^th^ April 2019).

Participants were prospectively recruited between 2^nd^ June, 2020 and 8^th^ July, 2020 and provided informed consent. Ethics approval was granted by the London-Brent Research Ethics Committee (REF 20/HRA/2510).

Electronic community prescription records were used to record community exacerbation events and electronic hospital records similarly for hospital exacerbations. The terms community and hospital exacerbation, rather than moderate or severe^2^, are used in in this study due to the known change in hospital admission criteria during the peak of the SARS-CoV-2 pandemic in England when hospital bed capacity was considered of critical importance. Community managed exacerbations were defined as those resulting in a prescription for oral corticosteroids and/or antibiotics but without hospital admission. Hospitalised exacerbations were defined as admissions to hospital with a recorded discharge diagnosis of Acute Exacerbation of COPD.

As an additional analysis a telephone survey was conducted to explore potential reasons for differences in exacerbation risk. Participants were asked to compare behavioural and emotional changes with their baseline “normal” state as a reference. Participants were asked to compare two discrete periods; (1) pre-lockdown, defined as the two weeks prior to “lockdown” (1^st^ March 2020 to 14^th^ March 2020) when participants were likely to be more aware of the threat of SARS-CoV-2 but restrictions had not yet been placed and (2) the “lockdown” itself (15^th^ March 2020 to 30^th^ April 2020). Self-reported behaviour included; medication adherence to their regular prescribed inhaled therapy, anxiety, self-reported change in activity levels, and social behaviour (self-isolation, shielding, visitors to the home, arrangements for shopping). Answers were captured with either a binary response (yes/no) or on a five-point Likert scale (see online supplement for full details of questionnaire used).

### Study Population

Participants were eligible if they had a confirmed diagnosis of COPD, under a specialist COPD clinic (Complex COPD clinic, Leicester, UK^11^), and able to provide informed verbal consent via English language telephone consultation. All patients had confirmed airflow obstruction. Patients were contacted sequentially from the research database held in our centre of patients who have previously consented to be contacted for research until this list was exhausted. The telephone call was made by either a nurse or doctor and participants gave informed consent verbally with this documented by the investigator due to the remote nature of the consultation.

Electronic GP and hospital healthcare records were used to capture new prescriptions for oral antibiotics or corticosteroids during the periods of interest, hospital admissions, as well as baseline characteristics, including latest spirometry. All spirometry had been performed at their previous clinic visit to Glenfield Hospital, Leicester to ERS/ATS standard^12^.

### Statistical Analysis

Baseline data were described as mean (standard deviation), or n (%) as appropriate. Paired data were compared using a paired t-test or signed-rank test for parametric data and non-parametric data respectively. Categorical data were compared using chi squared. Statistical analysis was performed using STATA 16 (StataCorp, USA).

From previous data from our COPD clinic we anticipated 0.8 exacerbations per patient in the observation period with a SD of 0.9. To detect a 25% difference in exacerbations within patients between 2019 and 2020 then 160 participants would be required (alpha=0.05, power 80%).

## RESULTS

160 patients were recruited with baseline characteristics outlined in table 1. 140 (88%) reported at least one exacerbation in the previous year, and the majority 103 (64%) reported at least two. 149 (93%) patients were prescribed triple inhaled therapy and 138 (86%) were classed as GOLD stage 3 or 4 airflow obstruction.

**Table 1:** Baseline Characteristics

|  | Mean (SD) |
| --- | --- |
| Age (years) | 67.3 (8.1) |
| Sex (male n, %) | 88 (55%) |
| FEV<sub>1</sub> (L) | 0.86 (0.12) |
| FEV<sub>1</sub> (% predicted) | 34 (13) |
| FVC (L) | 2.29 (0.87) |
| Smoking status (current/ex) | 31/129<br>(19%/81%) |
| Pack years | 52 (26) |
| Blood Eosinophil count $\geq 0.3 \times 10^9/L$ (n, %) | 58 (36%) |
| Spirometric GOLD Stage (n) |  |
| GOLD 1 | 10 (6%) |
| GOLD 2 | 12 (8%) |
| GOLD 3 | 72 (45%) |
| GOLD 4 | 66 (41%) |
| MRC Dyspnoea Score (n) |  |
| Grade $\leq 3$ | 34 (21%) |
| Grade 4 | 82 (51%) |
| Grade 5 | 44 (28%) |
| Moderate or severe AECOPD in past year (n) |  |
| 0 | 20 (13%) |
| 1 | 37 (23%) |
| $\geq 2$ | 103 (64%) |
| Hospitalisation for AECOPD in past year (n) | 99 |
| 0 | 99 (62%) |
| 1 | 38 (24%) |
| $\geq 2$ | 23 (14%) |
| Social circumstances (n) |  |
| Lives alone | 44 (28%) |
| Lives with partner/ spouse | 101 (63%) |
| Lives with children | 15 (9%) |

FEV<sub>1</sub>: Forced expiratory volume in one second, FVC: Forced Vital Capacity, AECOPD: acute exacerbation of COPD

### Number of exacerbations

In the first six weeks of the lock-down period (15^th^ March 20120 to 30^th^ April 2020) there were significantly more community and hospitalised exacerbations events compared with the same period in 2019 (126 vs 99 events, p=0.026). The number of exacerbations per patient are shown in figure 1a. Overall there was a similar proportion of patients who received treatment (34.4% in 2020 vs 33.8% in 2019) while those who received at least one course more likely to receive more separate courses of treatment (table 2).

**Table 2:** Number of participants suffering exacerbation events (prescriptions for antibiotics or corticosteroids or both in the community or admissions to hospital) separated by number of events recorded during 6 week reference period in each year.

|  | Number of events per participants | 2019 |  | 2020 |  |
| --- | --- | --- | --- | --- | --- |
|  |  | Number of Participant (n=160) | % | Number of Participants (n=160) | % |
| Community and Hospital Managed AECOPD Events | 0 | 106 | 66 | 105 | 66 |
|  | 1 | 14 | 9 | 10 | 6 |
|  | 2 | 35 | 22 | 21 | 13 |
|  | 3 | 5 | 3 | 22 | 14 |
|  | 4 | 0 | 0 | 2 | 1 |
| Community Managed AECOPD Events | 0 | 112 | 70 | 107 | 67 |
|  | 1 | 9 | 6 | 9 | 6 |
|  | 2 | 38 | 24 | 21 | 13 |
|  | 3 | 1 | 1 | 22 | 14 |
|  | 4 | 0 | 0 | 1 | 1 |
| Hospital Managed AECOPD Events | 0 | 150 | 94 | 155 | 97 |
|  | 1 | 9 | 6 | 5 | 3 |
|  | 2 | 1 | 1 | 0 | 0 |

**Figure 1:**
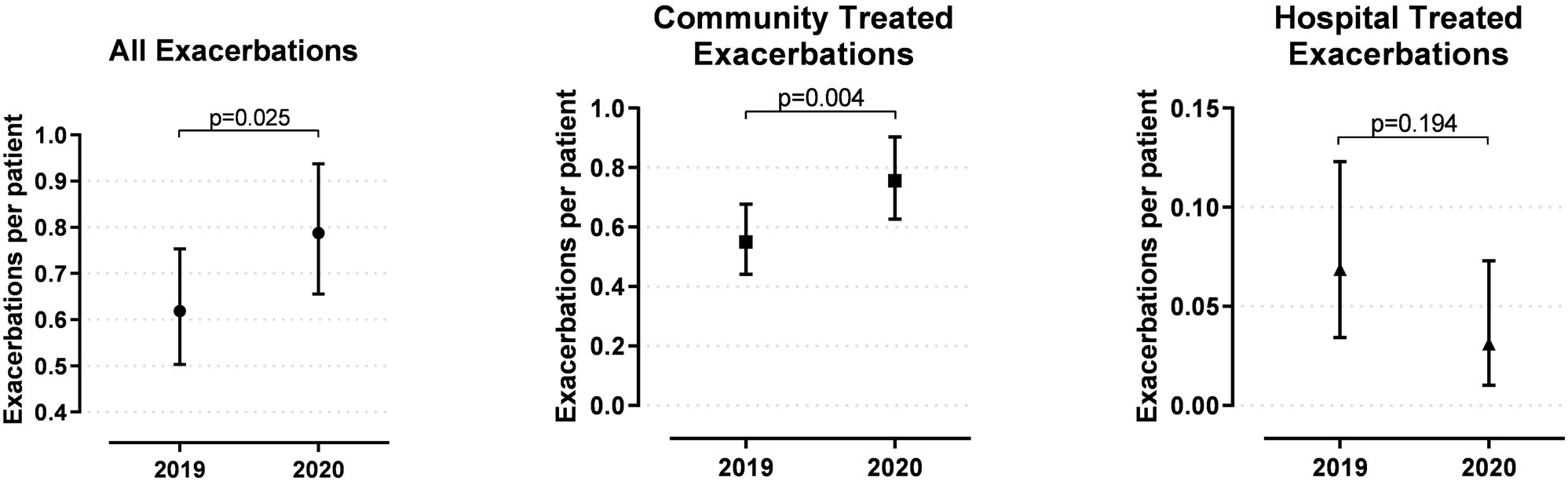
Number of exacerbations per patient between 15^th^ March and 30^th^ April in 2019 and 2020. (a) all community and hospitalised exacerbations (b) Community exacerbations only and (c) Hospitalised exacerbation only. Data shown are mean with 95% Poisson confidence interval.

Unsurprisingly, community managed events comprised the majority of exacerbations (209/224, 93%). For community exacerbations alone 121 events were noted in 2020 compared with 88 in 2019 (p=0.004) (figure 1b). There were 5 hospitalisations (n=5) due to AECOPD during the lockdown period in 2020, compared to 10 hospitalisations (n=9) in the same period in 2019 (figure 1c).

### Behaviour Pre-lockdown and Lockdown compared to Baseline

#### Medication Adherence

In the two weeks prior to lockdown 131 (83%) participants reported using their maintenance inhalers with the same frequency as they would during their stable state. 23 participants (14%) reporting increased use and 4 (2.5%) using less frequently than normal. During the lockdown period 42 (26%) participants reported increased use, 113 (71%) participants reported the same frequency of use and 4 (2.5%) reported using their regular inhaler less frequently than baseline (p<0.001) (figure 2a).

**Figure 2:**
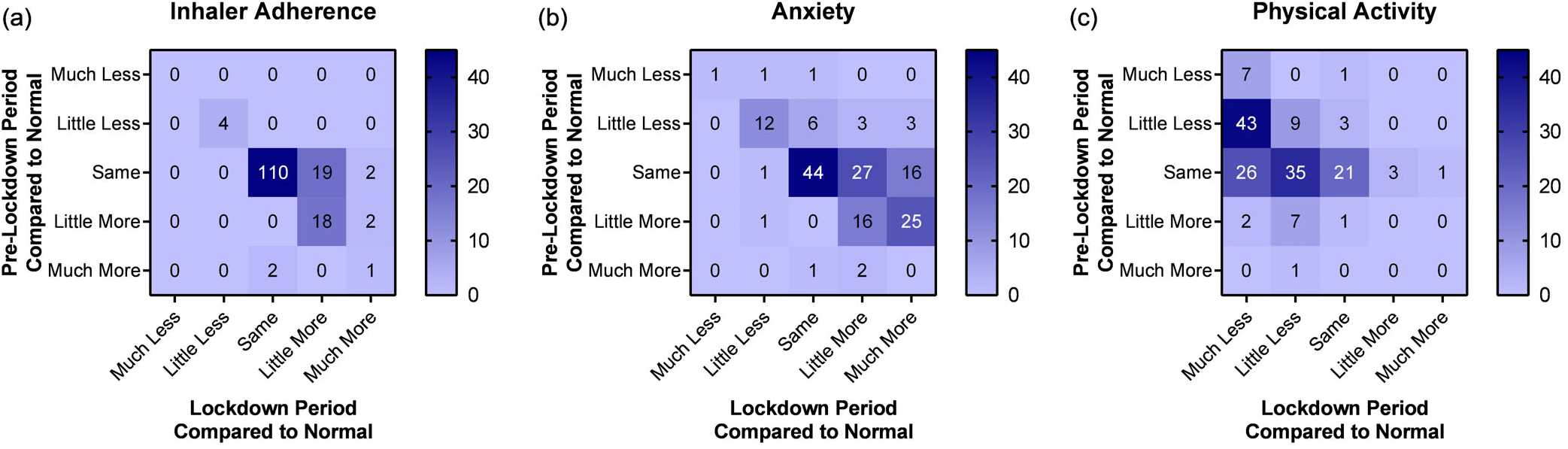
Changes in self-reported behaviour in the two weeks prior to lockdown (pre-lockdown) and during the first six weeks of lockdown (lockdown) compared to normal baseline for (a) Regular inhaler use (b) Anxiety (c) Physical Activity. Change in between pre-lockdown and lockdown for all groups p<0.001.

#### Anxiety

45 (28%) participants reported having more anxiety about their COPD than normal during the pre-lockdown period compared to baseline, of which 42/45 (93%) reported anxiety as a “little more” than baseline and 3/45 (7%) as “much more”. During the lockdown period 92 (58%) reported increased anxiety compared to normal (p<0.001), of which 48/92 (52%) were “a little more” anxious and 42/92 (48%) “much more” anxious (figure 2b).

Participants were also asked if they would avoid coming to hospital as an emergency during the pre-lockdown and lockdown periods due to fear of COVID-19. 64 (40%) reported they would have avoided doing so during the pre-lockdown and 90 (56%) reported they would avoid emergency hospital attendance during the lockdown period (p<0.001).

#### Physical activity and exercise

83 (54%) reported physical activity was unchanged compared to normal during the pre-lockdown period with 63 (40%) reporting reduced activity and 11 (7%) reporting increased activity levels. This contrasted sharply to the lockdown period where only 26 (16%) reported maintaining the same level of activity as normal while 52 (32.5%) reported slightly less and 78 (49%) reported doing a lot less physical activity than normal implying a significant decrease in activity levels (p<0.001). Only 4 (2%) reported increased physical activity levels (figure 2c). When asked about participation in a home exercise program, 50 (31%) patients and 56 (35%) patients reported participating in a home exercise program during the pre-lockdown and lockdown periods respectively.

#### Shopping behaviour

Participants were asked about shopping behaviour during the pre-lockdown and lockdown periods with a significant change being noted; during the pre-lockdown 89 (55.6%) reported going shopping themselves, while 33 (20.6%) reported that this was performed by someone who lives in the house with them and 38 (23.8%) reported it being completed by someone who does not live with them or being delivered to them. In contrast to this, during the lockdown only 11 (6.9%) reported that they still did their own shopping, with 37 (23.1%) having this task completed by someone living in their home and 112 (70%) reporting that it was done by someone who does not live in their home or delivered (pre-lockdown to lockdown, p<0.001).

#### Shielding and visitors

In the pre-lockdown period 142 (88.8%) participants reported continuing normal behaviour with only 16 (10%) shielding. Once lockdown started only 7 (4.4%) reported continuing normal behaviour while 127 (79.9%) reported that they were shielding (p<0.001).

During the pre-lockdown period 146 (91.3%) reported that they had visitors to their home compared to 31 (19.4%) during the lockdown (p<0.001).

## DISCUSSION

In this observational study a 38% increase in community managed exacerbation events during the COVID-19 lockdown in 2020 was seen compared to the same six-week period in 2019, as measured by primary care prescription records. The number of patients suffering an exacerbation was unchanged. Self-reported anxiety and inhaler adherence increased whereas PA was lower initially during the pre-lockdown period, but most pronounced during lockdown.

Severe exacerbations, as measured by hospital admissions, were seen within the cohort and represented 6% of all exacerbations. We observed a 50% decrease in hospital managed AECOPD events during the COVID-19 lockdown compared the same dates in 2019, though our study was insufficiently powered. A recent larger study, comparing hospital events, rather than individual patients, confirm our observations with a similar reduction in AECOPD admission rates^6^. This may represent an effect of the increased use of rescue medications in the community resulting in reduced need for hospital admissions, though other factors are also likely to have played a role.

This is the first report of the impact of the SARS-CoV-2 pandemic (and consequent societal lockdown) on objectively measured AECOPD rates. Our findings contrasts to reports of reduced AECOPD events during the lockdown with physicians and COPD nursing teams^5^. Interestingly we did not observe an increase in the proportion of patients requiring rescue medication, but an increase in the number of multiple courses. Possible explanations for these findings may result from either biological or behavioural differences. Patients who would normally have been admitted to hospital with an exacerbation may have been managed in the community during the pandemic because of a combination of fearfulness on the part of the patient about transmission risk in hospital and a desire on the part of healthcare teams to reserve hospital bed capacity to manage patients suffering with COVID-19 pneumonia. This Behavioural explanation appears plausible, particularly as there was increased access to healthcare services via telephone consultations and reduced physical access to clinicians^13^. National guidance, updated in 2018, recommended an “action plan” which includes oral corticosteroids and antibiotics to be self-administered in the event of an AECOPD^14^. Increased concerns on the part of clinicians about the risks of hospitalisation in a patient population perceived to be at greater risk from SARS-CoV-2 might have lowered thresholds for prescribing action plans. Patient concern that access to primary or secondary healthcare teams and pharmacies might be restricted might also have resulted in stockpiling behaviour during the pandemic^15^ with patients potentially requesting multiple “rescue packs” to store in case they were unable to obtain these later.

It is possible that the biological triggers for exacerbation events reduced for some patients because of lower respiratory virus transmission and air pollution during lockdown. However, our data suggest that this was outweighed by events driven by non-inflammatory causes, termed “pauci-inflammatory”^16^, which may be less responsive to oral antibiotics or corticosteroids^17^. This is supported by our observation that the majority of participants reported increasing anxiety about their COPD, particularly during the lockdown period. This would support the view that the most likely reason for the observed increase in exacerbations may be underpinned by behavioural change and concerns around COPD and emergency healthcare. It also highlights the need for potential psychological support in a vulnerable population, where anxiety and depression are common^18^.

In addition, it is clear from our data that adherence to shielding advice was widespread, likely reflecting a shared concern among patients about risks from COVID-19. Likewise, we observed an increase in self-reported inhaler compliance suggesting greater health concern and vigilance.

We also observed a greater dependence on others for day to day activities such as shopping and an overall reduction in physical activity among this cohort of patients with COPD that contrasts to that reported amongst the general population during the lockdown^19^. While this study did not directly assess the effect of this reduction in physical activity it raises the additional possibility that exacerbation events increased because of increased breathlessness and reduced resilience due to deconditioning and sarcopenia^20,21^. The longer term consequences of such altered activity behaviours remains to be seen but is of significant concern given the difficulty in providing timely and effective pulmonary rehabilitation in the context of the pandemic^22^.

At the time of writing, we are approaching winter in the northern hemisphere and no SARS-CoV-2 vaccine has yet been demonstrated to be safe and effective^23^, exacerbations of COPD are likely to increase with this season and result in increased hospitalisation. This study is a timely reminder that increased understanding of community prescribing practice and patient behaviour are important and may reveal effective tools in reducing morbidity and mortality in this group. Firstly, patients with COPD are going to require ongoing support and treatment, even if they are less likely to present to specialist or hospital services. Previous evidence has shown that pandemic influenza poses a significant risk to patients with COPD^24^ with the consequence that viral pandemics such as SARS-CoV-2 are likely to pose a similar risk. Developing robust and accessible systems to acutely review patients with COPD remotely to guide them in their use of rescue and preventer medication may reduce symptom burden, hospital admissions and unnecessary courses of potentially harmful oral corticosteroids and antibiotics. It is less likely that the increased number of moderate exacerbations recorded from our prescription data represent an increase in airway inflammation but rather a composite of increased anxiety and caution with the aim of preventing hospital admissions and the consequence that other, non-pharmacological, interventions may have been effective in managing these events^24^.

The conclusions drawn from this study are limited by both the relatively small sample size and the severity of the COPD seen in the cohort recruited. Though 160 patients has provided adequate power for statistically significant differences in community treated exacerbation and behavioural changes it has not been sufficiently large to detect changes in hospitalised events which would be better evaluated using larger datasets. In addition to this the cohort had established COPD, under a specialist secondary care clinic, so results may not be applicable to those with milder disease, and less frequent exacerbations. Finally, patients recruited needed to be alive during the period of recruitment in May and June 2020, meaning that there may be survivor bias compared to those that died in 2019 and during the peak of the pandemic.

In summary, this study revealed an increase in treatment for community treated AECOPD events among patients with severe COPD during the SAR-CoV-2 lockdown. This finding was unexpected but may be explained by factors such as anxiety, which was increased in our patient cohort. Significant behaviour changes including reduced physical activity, adherence to shielding advice and increased inhaler compliance.

## Data Availability

All data requests should be submitted to the corresponding author for consideration. Access to anonymised data may be granted following review.

## Acknowledgements

We wish to acknowledge the work of the following members of the NIHR Leicester BRC Respiratory team and University Hospitals of Leicester: Sarah Parker, Jo Finch, Sarah Glover, Tracy Thorn, Bo Zhao, and Adrian Manise.

